# Relating *in vivo* RSV infection kinetics to host infectiousness in different age groups

**DOI:** 10.1101/2024.11.14.24317347

**Authors:** Ke Li, Louis J. Bont, Daniel M. Weinberger, Virginia E. Pitzer

## Abstract

Respiratory syncytial virus (RSV) infections are a major public health concern for pediatric populations and older adults. Viral kinetics, the dynamic processes of viral infection within an individual over time, vary across different populations. However, RSV transmission in different age groups is incompletely understood from the perspective of individual-level viral kinetics. To explore how individual viral kinetics can be related to RSV transmission, we first fitted a mathematical model to longitudinal viral kinetic data from 53 individuals in pediatric, adult, and elderly age groups using a hierarchical Bayesian framework to estimate important viral kinetic parameters. Using a probabilistic model, we then related the within-host viral load to the probability of transmission for each age group. We found that children had higher peak viral loads and longer shedding periods compared to other age groups, suggesting a higher transmission probability in children over the infectious period. We validated our findings by comparing the estimated secondary attack rate across different age groups to empirical estimates from household transmission studies. Our work highlights the importance of age-specific considerations in understanding and managing RSV infections, suggesting that age-targeted interventions will be more effective in controlling RSV transmission.

**Summary:** We utilized within-host viral load kinetics data to infer the transmission potential of RSV infection across different age groups, revealing the highest transmission probability in the pediatric group.

## Introduction

Respiratory syncytial virus (RSV) infections pose a significant public health threat to pediatric populations, older adults, and immunocompromised individuals [1]. In 2019, an estimated 100,000 deaths of children under the age of 5 were attributed to RSV globally [2]. Understanding transmission risks is of great importance to reducing the burden of RSV infections. Different age groups exhibit varying levels of susceptibility to RSV infection [3–7], which may necessitate age-specific strategies for prevention and control. However, relatively little is known about the clinical parameters affecting viral load of RSV across different demographic groups, although this has been shown to affect disease severity and transmission for other pathogens [8,9]. In this study, we aim to explore the quantitative relationship between RSV viral load and infectiousness in different age groups.

Mathematical models that depict viral dynamics have been used to study the relationship between viral load and infectiousness for different pathogens [8,10,11]. These models have advantages in being able to fit data and estimate important virological parameters. Previous modeling work utilized RSV viral load data from different experimental settings (e.g., *in vitro* and *in vivo*) to estimate kinetic parameters for RSV infection [12]. However, this work left room for methodological improvement. The study did not quantify the uncertainty in the estimated parameters, which limits the ability to draw reliable conclusions on differences between age groups based solely on point estimates and to examine the implications for viral transmission.

In this work, we first constructed a mathematical model for the viral load kinetics of RSV infection. We fitted the model to longitudinal viral kinetic data from 53 individuals across three age groups simultaneously in a hierarchical Bayesian framework to estimate important parameters, including viral shedding period and viral growth rate. We then associated viral load data measured by quantitative reverse transcriptase-polymerase chain reaction (qPCR) with viral load data measured by viral culture assays to explore the relationship between total viral load and the number of infectious virus particles during RSV infection. Using this relationship and a probabilistic model, we further predicted the transmission probability of RSV infections among pediatric, adult, and elderly age groups.

## Results

### Dynamics of RSV infection captured by a mathematical model

To explore whether within-host RSV kinetics vary among different age groups, we analyzed four different sets of longitudinal viral load data from pediatric [13,14], adult [15], and elderly groups [12,16] (see **Data sources** for a detailed data description). In total, viral load data from 53 individuals were studied (pediatric group: 24; adult group: 7; elderly group: 22). The data from the pediatric and elderly groups were obtained from observational studies, whereas data from the adult group were obtained from a human challenge study. Therefore, the infection time was unknown for the pediatric and elderly individuals and had to be estimated.

To estimate key parameters of viral kinetics, we fitted the data simultaneously using an empirical mathematical model (**Eq. (1)** in **Methods**) that captures the typical viral dynamics of acute respiratory infections using a hierarchical Bayesian framework (see **Methods** for details). This allows us to estimate the magnitude and timing of the peak viral load, as well as the rate of growth and decline of the viral load. Based on the median estimates of the model parameters, we showed that our model could successfully replicate the viral kinetics observed in the three age groups, capturing both the growth phase, the peak, and the decline phase of the viral kinetics (black curves, **Fig. 1 and Fig. S1-S3**). We also randomly simulated 1000 viral load trajectories based on the joint posterior distribution of the estimated parameters (gray curves, **Fig. 1**), all of which closely matched the trend of the data. For each individual in the pediatric and elderly groups, we also estimated the time of symptom onset relative to time of infection. This is because the infection time was unknown for the groups. We found that the time of symptom onset relative to the time of infection was not notably different between the two age groups, with means of 3.54 days (95% PI: [0.94, 6.60]) and 3.50 days (95% PI: [1, 6.32]) in the pediatric and elderly groups, respectively (**Fig. S4**).

**Figure 1.**
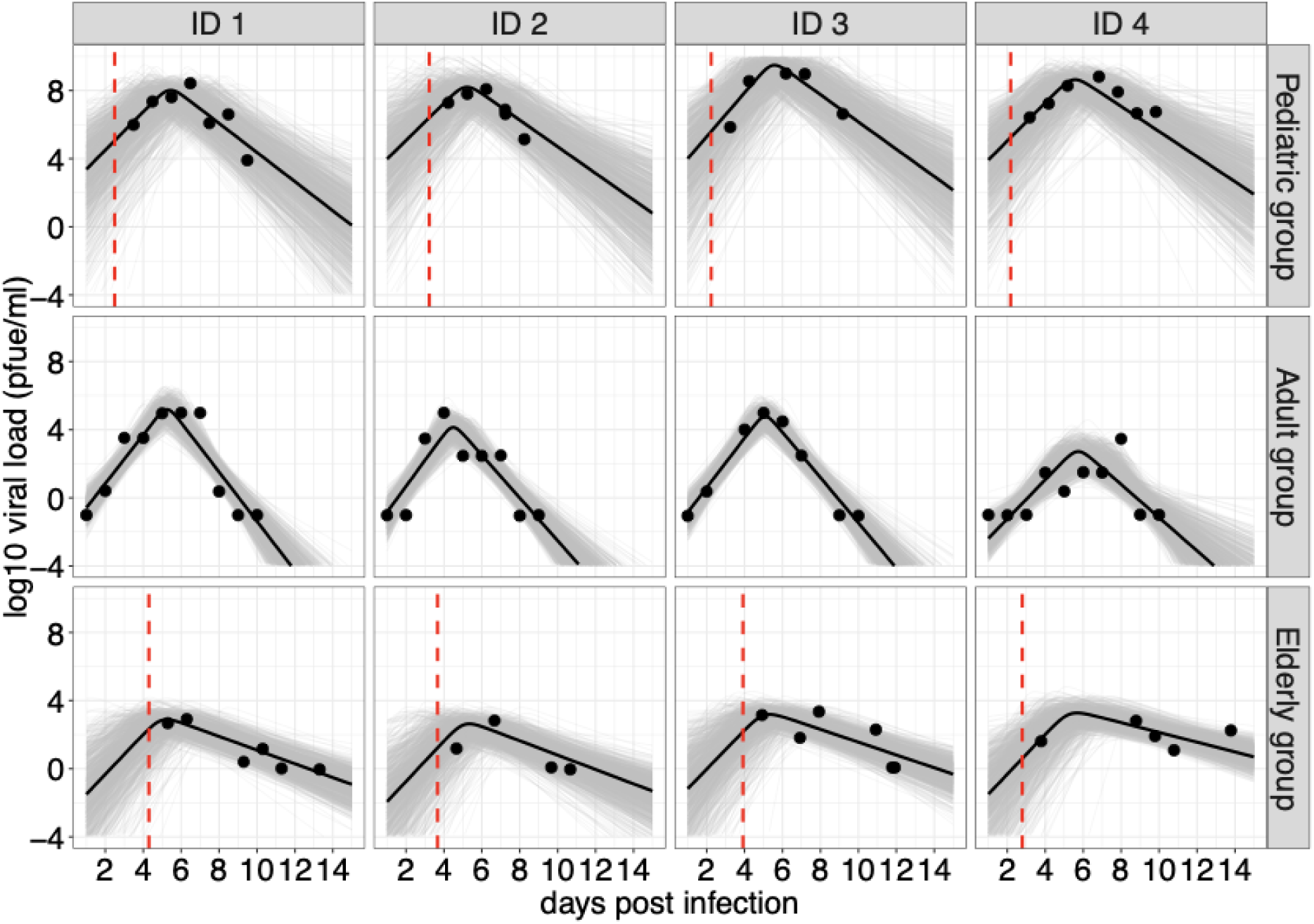
Model fits to viral kinetic data of RSV infections in different age groups. Data are presented with solid circles, and solid lines are median predictions of viral load. The dashed red lines indicate the median estimates of the time of symptom onset relative to time of infection for the pediatric and elderly groups. Note that the median predictions of viral load are calculated using estimated parameters for each individual. Gray curves are 1000 simulated viral load trajectories based on the joint posterior distribution. Four individuals from each age group were selected for presentation; results for all individuals are provided in **Fig. S1-S3**.

### Viral kinetics vary among age groups

Having observed that our model can successfully reproduce the viral kinetics data, we next examined the differences in model parameters among the age groups. We observed that the pediatric group exhibited a significantly higher peak viral load (*p* < 0.01, Wilcoxon Rank-Sum Test), with a median of 5.84 (a 95% credible interval (CI): [5.12, 6.80]) log10(plaque-forming equivalent units/ml) (PFUe/ml), compared to the adult group (median = 4.14, 95% CI: [3.37, 5.01]) and the elderly group (median = 2.96, 95% CI: [2.63, 3.35]) (**Fig. 2A**). We also compared the posterior distribution of the variance of peak viral load and found that variance was largest for the pediatric group (median variance = 1.25 log10 viral copies (**Fig. S5A**). This suggests significant heterogeneity in peak viral load within this group, which may be associated with varying conditions (e.g., previous infections) among different pediatric individuals. Furthermore, we found no difference in the time to peak viral load among the adult and elderly groups (*p* > 0.05, Wilcoxon Rank-Sum Test, **Fig. 2B**), with the median estimated to be approximately 5 days. In contrast, we observed that the pediatric group had an early peak time compared to the other age groups with a median of 3.09 (95% CI: [1.55, 5.18]) days. Similarly, we observed comparable growth rates of viral load between the adult and elderly groups, with medians of 3.21 (95% CI: [2.59, 4.03]) and 3.15 (95% CI: [1.15, 6.27]) log10(PFUe/ml) per day, respectively, while the growth rate was slightly higher in the pediatric group (median = 2.82, 95% CI: [0.86, 6.63]) (**Fig. 2C**). The adult group also had the fastest rate of viral clearance (median = 2.71 (95% CI: [2.09, 3.63]) log10(PFUe/ml) per day), whereas the pediatric and elderly group had the slower rate of viral clearance, with medians of 1.01 (95% CI: [0.83, 1.26]) and 0.87 (95% CI: [0.67, 1.18]), respectively (**Fig. 2D**). We did not observe significant heterogeneity within each age group, nor a significant difference in variance between age groups for these parameters (**Fig. S5B-D**).

**Figure 2.**
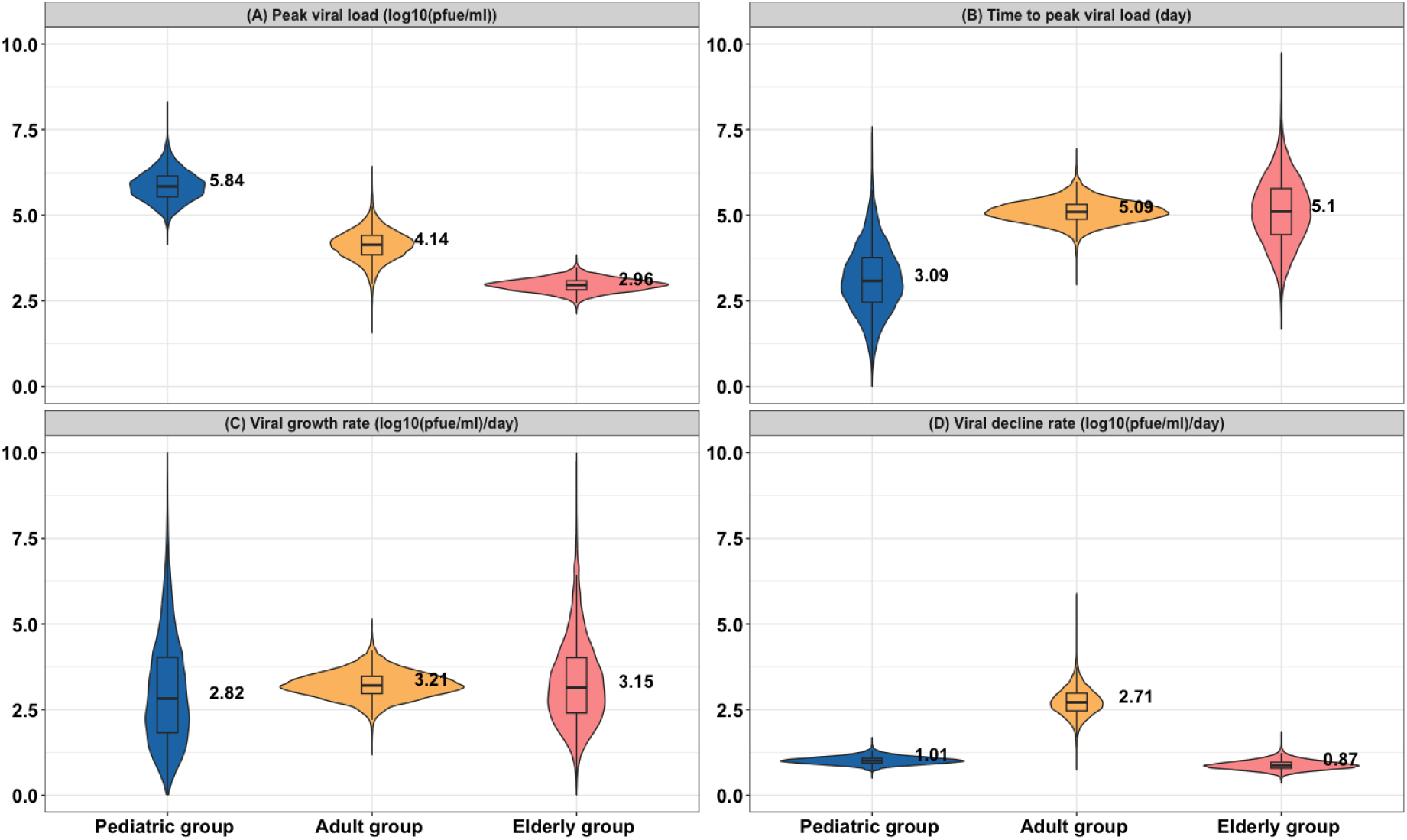
Posterior distributions of key model parameters. 12,500 samples are drawn from the population-level posterior distributions of **(A)** peak viral load (log10 PFUe/ml), **(B)** time to peak viral load (days), **(C)** the growth rate of viral load (log10 PFUe/ml/day), and **(D)** the decline rate of viral load (log10 PFUe/ml/day) in the three age groups. Dashed lines indicated the median value of the posterior distributions.

We hypothesized that the significant differences in peak viral load among the groups could depend upon the initial viral load (i.e., viral load at infection time *t* = 0). Based on the posterior samples, we calculated the initial viral load for each group and found that the pediatric group had the highest initial values compared to the other groups, while no difference was observed between the elderly and adult groups (**Fig. S6**). We further looked into and calculated the initial viral load based on estimated individual-level heterogeneity and found that among the infected individuals, pediatric individuals had higher initial viral load compared to the individuals in other age groups (**Fig. S7**). This suggests that the high peak viral load in the pediatric group is related to higher initial viral load, whereas the higher peak viral load in the adult group compared to the elderly group appears to be due to a faster growth rate (**Fig. 2C**).

### Comparison of viral shedding periods measured by qPCR and cell culture methods

Having revealed the difference of viral kinetics among age groups, we next sought to explore how within-host dynamics relate to viral transmission at the population level. To do this, we applied a probabilistic model from [8] to describe various steps in viral transmission, from within-host viral shedding in an infector to the establishment of infection in a recipient. Here, infectiousness is defined as the probability that an infector produces one or more infectious viral particles and transmits them to a recipient, leading to a successful infection.

We first quantified the relationship between the number of infectious viruses and the measured total viral load using quantitative reverse transcriptase-polymerase chain reaction (qPCR). By utilizing the data from human challenge studies [15,17] (see **Data sources**), we found that saturation models (**Fig. 3A** and **3B**) best described the relationship compared to linear or power-law models (**Fig. S8**), showing that the level of infectious viruses, measured by a culture technique, increases sublinearly with increases in total viral load measured by qPCR. Notice that the range of qPCR values differs between the studies because the detection limits are different.

**Figure 3.**
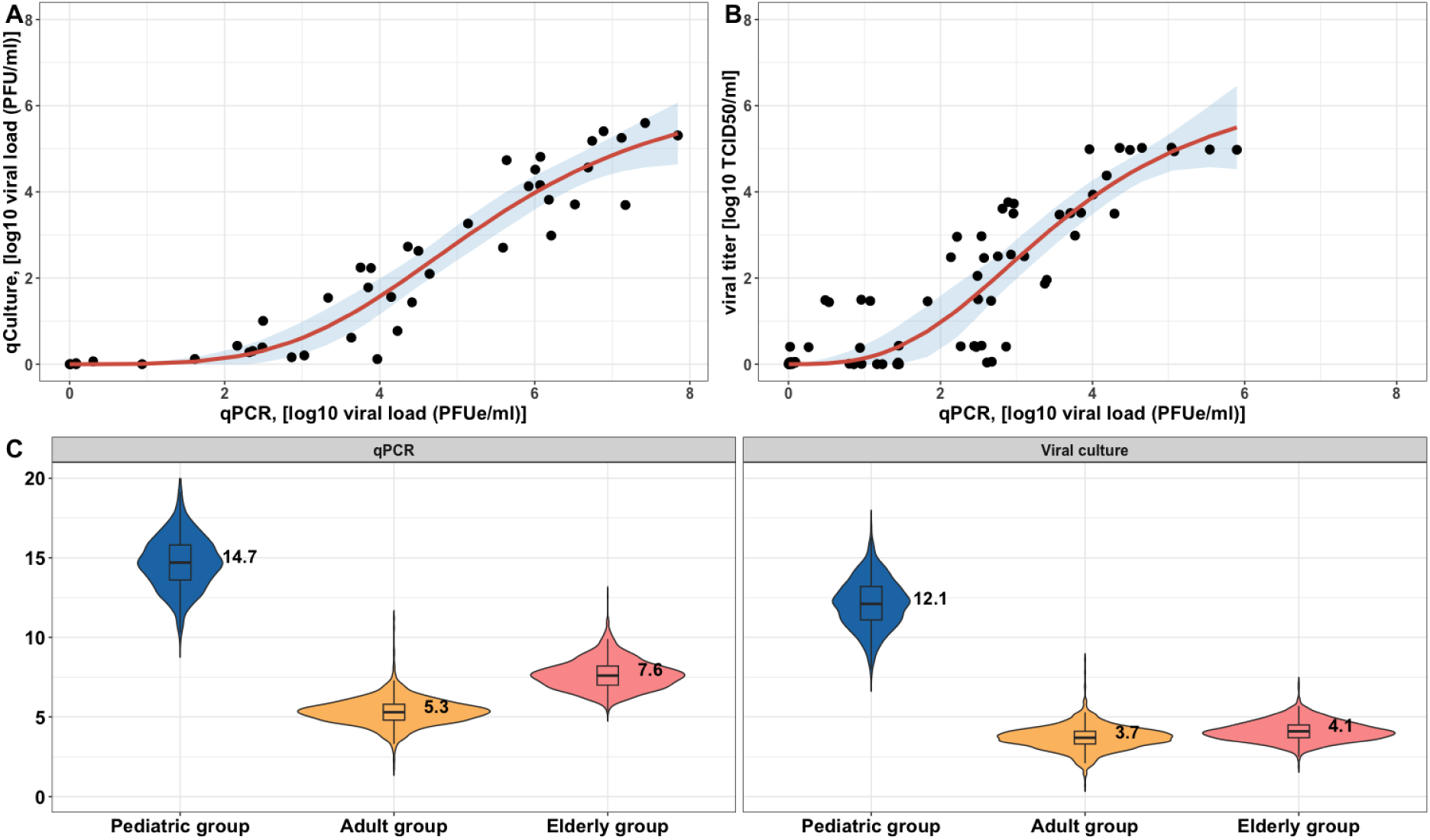
The characterization of the relationship between the viral concentration measured by qPCR and viral culture. The best fit of using a saturation model (in a form of 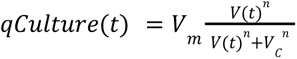, where *V*(*t*) is the viral load measured by qPCR) to the data from (**A**) [17], *V_m_* = 6.59 log10(PFU/ml), *n* = 3.91, *V_c_* = 5.39 log10(PFUe/ml) and (**B**) [15], *V_m_* = 6.78 log10 (TCID50/ml), *n* = 3, *V_c_* = 3.64 log10(PFUe/ml). Black circles are data points. Mean predictions are given by the red curves, and the shaded areas indicate a 95% confidence interval. 12,500 samples are drawn from the population-level posterior distribution to calculate **(C)** viral shedding period (in days) (defined as the duration when viral load (measured by qPCR or viral culture) is above 10 PFUe/ml or PFU/ml) for different age groups.

Based on the relationship between the total viral load measured by qPCR and the level of infectious viruses, we then predicted the expected viral shedding period for each age group using the two measurements. If we set the limit of detection to 10^1^ PFUe/ml of viral load measured by qPCR, then the pediatric group had the longest expected shedding period with a median of 14.7 days, compared to the adult group (median = 5.3 days) and the elderly group (median = 7.8 days) (**Fig. 3C**). The duration of expected RSV shedding decreased when the viral load was measured by viral culture with the limit of detection of 10^1^ PFU/ml. The limit of detection was set based on [15]. We observed that the median estimates of the duration were 12.1 days, 3.7 days, and 4.1 days for the three age groups, respectively.

### Modeling transmission potential of RSV infection in different age groups

Following the characterization of the relationship between infectious virus and total viral load, we next modeled the probability of establishing an infection in a recipient (see **Fig. S9** for an illustrative diagram). Similar to the probabilistic model described in [8], we assumed that the number of infectious virions shed by an infector at time *t* during the shedding period is *v*(*t*). On average, the number of infectious viruses reaching a recipient during a contact at time *t* is a random variable *X_v_* that is Poisson distributed with mean *v*(*t*), such that *X_v_* (*t*)∼ *Poisson*(*v*(*t*)). We further assumed that each viral particle shed by an infected individual has a probability *p_v_* to establish infection in a recipient during a household contact.

Taken together, the distribution of *X_v_* viruses that reach and establish an infection in a recipient at time *t* follows a Poisson distribution with mean and variance 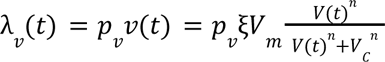, where the parameter ξ accounts for the number of infectious particles derived from the measured viral concentration (i.e., PFU/ml). We assumed ξ varies among age groups to reflect the differences in lung capacity, which could affect the production and spread of respiratory droplets.

We computed the probability of a single infectious virus establishing infection (*p_v_* ) based on empirical estimates of the secondary attack rate (SAR) from RSV household transmission studies [18–20]. The probability of a contact in a household becoming infected during the viral shedding period (*P_inf_*) can be expressed as 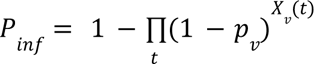 (see **Methods** for details). Taking into account the cumulative viral load during the viral shedding period (**Eq. (2)** in **Methods**), we calculated *p_v_* based on the SAR for pediatric individuals (**Table 1**). We calibrated ξ to account for the difference in respiratory droplets spread between pediatric individuals and other groups, based on the SAR for the adult group from [18]. By comparing the predicted SAR with the empirical estimates, we found that the predicted SAR in the adult group was comparable to the empirical estimates from [19], but it was underestimated when compared with the SARs from [20]. This discrepancy may be due to the small sample size.

**Table 1.**
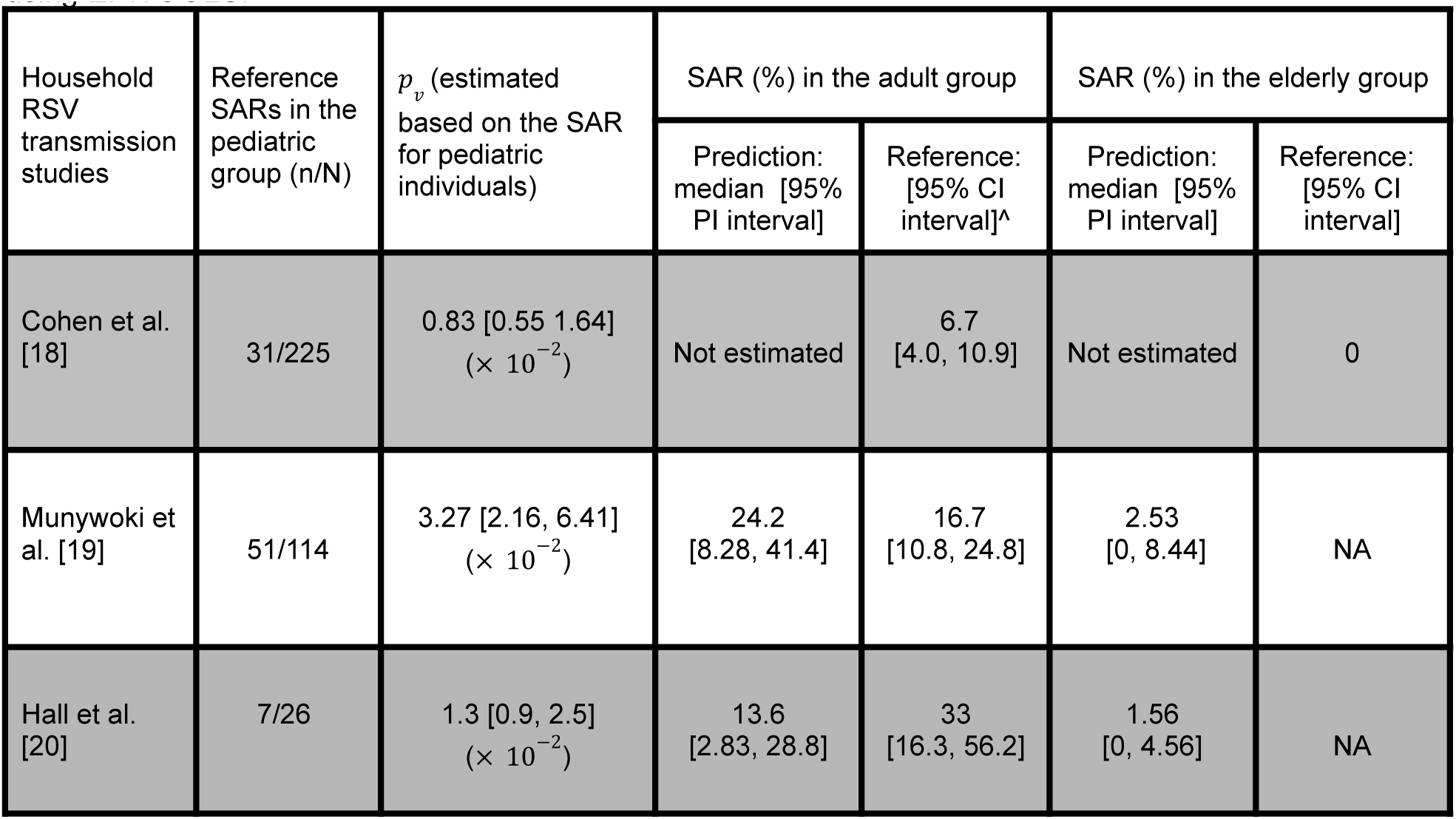
Estimates of infection probability and secondary attack rates. The secondary attack rate pediatric individuals (1-4 years old) were used to compute the infection probability for each study. The parameter ξ was calibrated only based on the median estimates of *p_v_* and the SAR in the adult group from [18] (median = 4.57 with a 95% PI [2.03, 23.36]). Using the median estimates of ξ, the SARs for the adult group (15-49 years old) in [19] and in (17-45 years old) [20] were computed. The SARs for the elderly group (>65 years old) were not available. ^The 95% confidence interval (CI) was estimated using EPITOOLS.

| Household RSV transmission studies | Reference SARs in the pediatric group (n/N) | $p_v$ (estimated based on the SAR for pediatric individuals) | SAR (%) in the adult group | | SAR (%) in the elderly group | |
| --- | --- | --- | --- | --- | --- | --- |
|  |  |  | Prediction: median [95% PI interval] | Reference: [95% CI interval]^ | Prediction: median [95% PI interval] | Reference: [95% CI interval] |
| Cohen et al. [18] | 31/225 | 0.83 [0.55 1.64]<br>($\times 10^{-2}$) | Not estimated | 6.7 [4.0, 10.9] | Not estimated | 0 |
| Munywoki et al. [19] | 51/114 | 3.27 [2.16, 6.41]<br>($\times 10^{-2}$) | 24.2 [8.28, 41.4] | 16.7 [10.8, 24.8] | 2.53 [0, 8.44] | NA |
| Hall et al. [20] | 7/26 | 1.3 [0.9, 2.5]<br>($\times 10^{-2}$) | 13.6 [2.83, 28.8] | 33 [16.3, 56.2] | 1.56 [0, 4.56] | NA |

We then simulated 12,500 viral load trajectories using posterior samples and predicted the probability of transmission during infection in different age groups, given the median estimates of infection probabilities from [18] (**Fig. 4A**). The probability of one or more viruses successfully transmitting from an infector and establishing an infection in a recipient during a contact at time *t* is given by *p*(*t*) = 1 − *exp*(− λ_*v*_ (*t*)). We found that the pediatric group exhibited the highest predicted transmission probabilities throughout the infections compared to the other two groups. Compared to the elderly group, the adult group had a higher predicted probability of transmission during infections. This was due to the higher viral load in the adult group. In addition, we predicted that the highest transmission would occur during the time of symptom onset in the pediatric group (shaded areas in **Fig. 4A**). Note that the results showed the expected difference in probability of infection that would be expected from different viral loads by age groups. We also observed qualitatively similar results given the median estimates of infection probability from [19,20] (**Fig. S10**). We also showed that the rate of presymptomatic transmissions (**Eq. (4)** in **Methods**) was low in the pediatric and elderly groups (**Fig. 4B**). The median value of the predicted fractions of presymptomatic transmissions was 2.6% (95% PI: [0, 81.9%]) in the pediatric group, whereas a median value of 7.26% (95%PI: [0, 47.5%]) of presymptomatic transmissions occurred in the elderly group. We did not calculate the fractions for the adult group, as the time of symptom onset was not estimated for this group.

**Figure 4.**
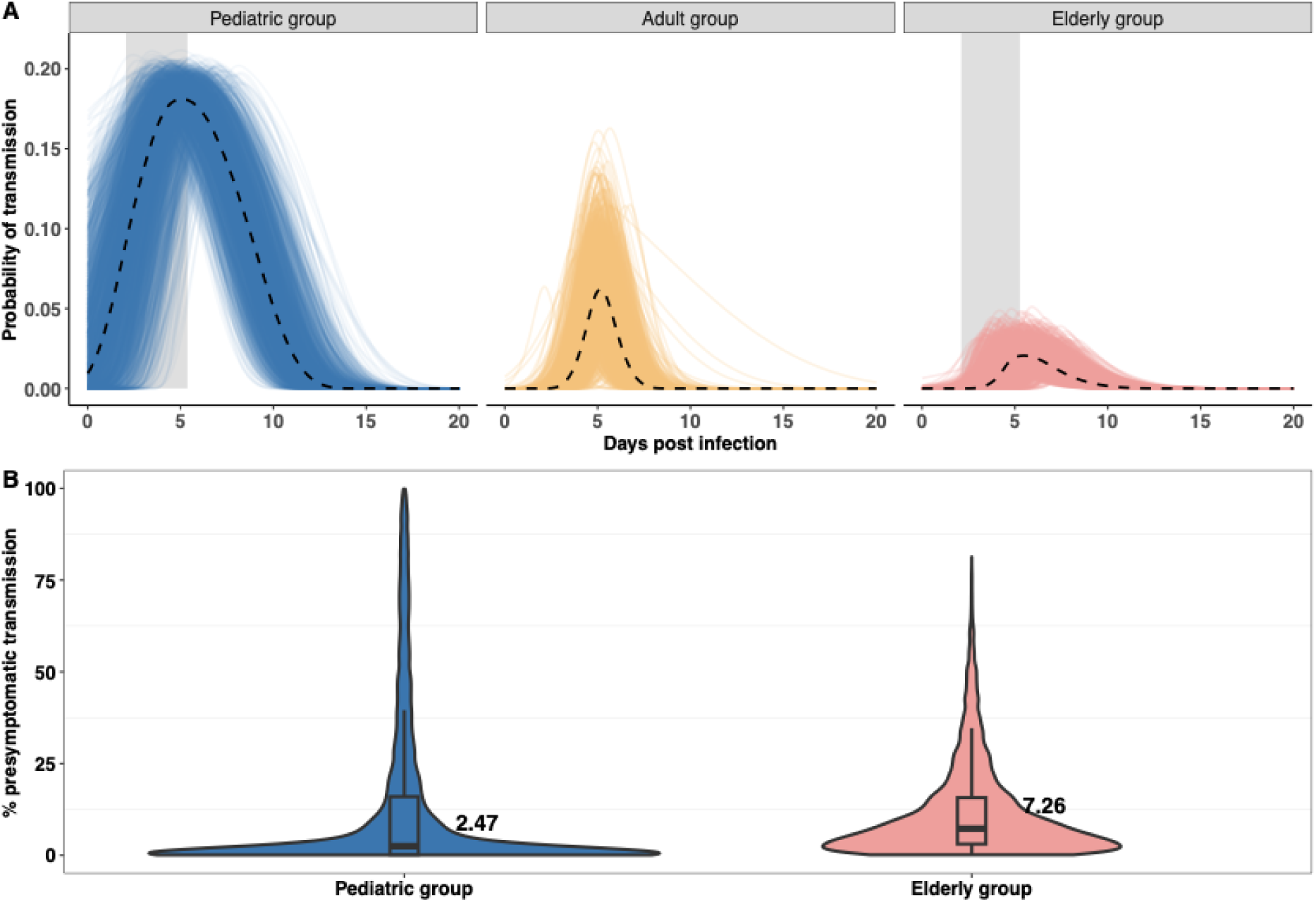
Probability of transmission and predicted presymptomatic fractions in different age groups. **(A)** 12,500 population-level samples are drawn from the posterior distribution to calculate the probability of transmission as time-series in different age groups, given the median estimates of infection probability from [18–20]. Dashed lines indicate the median trajectory, and shaded areas indicate estimated incubation period in the pediatric and elderly group. **(B)** The percentage of presymptomatic transmissions for the pediatric group and the elderly group based on the 12,500 population-level posterior samples.

## Discussion

In this study, we applied a mathematical model to describe the within-host viral kinetics of RSV infections among individuals from pediatric, adult, and elderly age groups. We estimated viral kinetic parameters in a hierarchical Bayesian framework. The statistical method allowed us to account for and explain underlying heterogeneity in the viral load data at both the population and individual levels, providing robust estimates of viral kinetics parameters across the different age groups. We derived posterior distributions for key parameters such as the peak viral load, growth rate, and clearance rate, which were then used to assess differences in viral dynamics among different age groups. Our analysis revealed and quantified distinct patterns in viral load trajectories, with the pediatric group exhibiting a higher initial and peak viral load compared to the adult and elderly groups, and this was supported by a prospective cohort study [21] and others [22–24]. In contrast, elderly individuals had the lowest peak viral loads during infection but also exhibited the slowest viral clearance rates, suggesting a reduced ability to mount an effective immune response to RSV [25–27]. Based on the viral load data, we also estimated the incubation period in the pediatric and elderly groups and found no difference between them, with medians of 3.95 and 3.41 days, respectively. This differs from studies estimating the incubation period of RSV at 4.4 days (95% confidence interval: [3.9-4.9]) [28] and reporting a range of 2 to 8 days depending on age [29]. The variations could be attributed to different estimation methods.

While qPCR provides a measure of the total viral load [30], including both infectious and non-infectious particles, viral culture specifically quantifies the infectious viral particles [31]. Characterization of the relationship between the two measurements is crucial for understanding the progression of viral production and viral transmission. We examined the relationship between the level of infectious viruses and total viral load, measured by viral culture and qPCR, respectively, during RSV infection. This was achieved by fitting different forms of mathematical model to both viral load data. We found that the number of infectious virions followed a sublinear increase with total viral load. A similar finding was reported in a recent SARS-CoV-2 study [8]. In our study, the relationship between infectious virus and total viral load was derived based on human challenge studies in adult volunteers and may be different for other age groups. Future clinical studies should measure both total and infectious viral loads across various age groups.

Quantifying the infectious period and the duration of viral shedding are important for future models to understand RSV transmission. A strength of our study is the ability to estimate and compare the duration of RSV shedding using two different measurements of viral load, qPCR and viral culture. Our presented durations of RSV shedding are comparable to a recently published estimate of 11.3 days for the pediatric group (i.e., age <1 year old), 5.5 days for the adult group (i.e., age 13-64 years old), and 7.2 days for the elderly group (i.e., age >65 years old) [18]. We also found that the estimates of RSV shedding were longer when using qPCR compared to viral culture, although the pediatric group had the longest shedding duration irrespective of different methods of measurement. If we set the limit of detection to 10 PFU/ml based on [15], we noted that the estimated duration of viral shedding in the adult and elderly groups was largely reduced when changing the measurement method from qPCR to viral culture, implying less infectious viruses were produced in elderly individuals.

Estimating the secondary attack rate (SAR) is important for understanding RSV transmissibility and informing effective public health interventions. Previous household transmission studies have shown that SAR differs by age group [18–20], indicating that the risk of RSV transmission varies across different age groups. Our study demonstrated that viral load data could provide additional insights for understanding differences in RSV transmission. We computed the SARs based on the longitudinal viral load data for different age groups through rigorous mathematical and statistical formulations, and our results are comparable with empirical estimates. This validates our assumption that age-specific viral kinetics contribute to the difference in transmission risks across different age groups. Our study also suggests that combining viral load kinetic data with observational data may help more accurate estimation of SARs. Observed SARs might be biased because they attribute all “secondary” cases among household members to the first symptomatic individual (i.e., the index case) and do not account for the additional risk of infection from external sources or other infected household members. Keeping a record of viral kinetic data could help differentiate between infections caused by the index case and those resulting from external sources or other household members, providing a better estimation of SARs.

By constructing a probabilistic model capturing viral transmission and infection from infectors to recipients, our model predicted that pediatric individuals would show the highest transmission potential during their infection due to expected higher viral load, while adults and elderly individuals would have a lower transmission probability. This implies that children play a significant role in the spread of RSV, as also suggested by other epidemiological studies [18,32,33]. Here, the probability of transmission reflects only the infectiousness of an individual based on the predicted viral load and does not account for the frequency of contacts. Assuming that children can have more frequent contacts than those in other age groups, the transmission probability may be even higher in the pediatric group [34,35].

Recognizing and addressing presymptomatic transmission is crucial for effective disease control and prevention efforts [36]. We predicted that the rate of presymptomatic transmission was low in both children and elderly individuals. Compared to SARS-CoV-2 infections, where presymptomatic transmission accounted for a considerable portion of COVID-19 spread [37], we estimated here that RSV transmission is primarily driven by symptomatic individuals in the pediatric age group. We hypothesize that this is because RSV is more likely to cause lower respiratory tract infection in children [38], and infected individuals tend to have more severe symptoms while transmitting the virus.

There are limitations to our study. First, study settings were different in the adult group compared to the other two groups, as the adult group involved a human challenge study. In the experimental challenge study, individuals may be inoculated with higher doses than those experienced in natural infections, thereby influencing overall viral kinetics. Thus, the model estimates were less comparable in the adult group compared to the pediatric and elderly groups. Also note that we could not account for the lack of standardization of qPCRs between different studies, and the absolute qPCR values may not be comparable across age groups. Nevertheless, the viral load data for pediatric individuals, although obtained from two different studies, was determined using the same plaque assay method [39]. Second, we acknowledge that fewer data points were collected during the viral load growth period compared to the viral decay period among most individuals in the pediatric and elderly groups. This explains the large credible intervals for the estimated viral growth rate for the pediatric and elderly groups. Further work may be needed to extend our analysis to other datasets with more data points. We also note that we only were able to identify viral load data from 53 individuals, and additional data may be needed to draw more robust conclusions. In addition, the virological data were collected from pediatric and elderly patients who presented at hospitals and were likely to have more severe illnesses. Therefore, this could affect parameter estimates, considering the correlation between viral load and clinical symptoms [17].

Overall, our study defines viral load dynamics of RSV infections and highlights the importance of viral load dynamic data, measured by both qPCR and viral culture, in understanding RSV infections within different demographic groups. We argue that viral kinetic data provides insights into disease progression in natural settings, with potential to improve understanding of the spread of RSV and to test the effectiveness of clinical interventions such as antiviral therapeutics. Our model can be extended to study when and how to intervene in viral replication, particularly in children and elderly individuals, and to evaluate disease outcomes. Given the high viral loads in pediatric patients, early clinical interventions could reduce viral replication and transmission potential. Our work also establishes a mathematical framework to better understand the transmission potential of RSV infections among different age groups by utilizing individual-level viral load kinetic data. This approach enhances our comprehension of RSV transmission dynamics and improves the accuracy of epidemiological models. In the future, our work could be used to inform a multiscale mathematical model that combines within-host virological data and population-level incidence information, incorporating transmission heterogeneity at the group level, to predict RSV transmission and to inform and evaluate public health interventions.

## Methods

### Data sources

We conducted a review of literature, searching for viral load kinetic data by using keywords such as “*RSV infection*”, “*longitudinal data*”, “*viral load*”, “*RSV human challenge study*” and “*RSV viral dynamics”*. We filtered for studies with viral load kinetic data that included more than two data points for each individual. We analyzed four different sets of longitudinal viral load data from pediatric [13,14], adult [15], and elderly groups [16]. The *in vivo* viral load kinetic data were extracted using <u>WebPlotDigitizer</u> (version 5).

For the pediatric group, previously healthy children aged under 2 years were prospectively enrolled if they had RSV detected from respiratory secretions within the past 48 hours. Nasal aspirates were obtained quantitatively at enrollment using a standardized technique [39]. A reverse transcriptase–PCR was performed on cultured patient RSV isolates grown in HEp-2 cells.

For the adult group [15], healthy adults, ages 21 to 50 years, were inoculated with RSV A2. Infection was established by nasal inoculation of virus at a dose of 10 ^4.7^ 50% tissue culture infective doses (TCID50). Subjects underwent nasal washes on day 0 (prior to challenge), days 1 through 12, and day 28 postinfection. Quantitative RT-PCR was performed on all samples.

For the elderly group [16], we analyzed the viral load data of patients aged greater than 65 years who presented with influenza-like symptoms at the hospital. Viral titer measurements were taken on the day patients presented with symptoms and for several days after. RSV infection was diagnosed using real-time RT-PCR performed on initial nasal swab and sputum specimens.

To quantify the relationship between the number of infectious viruses and the measured total viral load using quantitative reverse transcriptase-polymerase chain reaction (qPCR), we utilized data comparing a qPCR assay with a culture technique to quantitatively assess viral load in adults challenged with RSV [15,17]. In total, we extracted viral load data, measured by both qPCR and viral culture assays, from 53 volunteers for further analysis. We examined and fitted different models to the data describing the relationship between viral load measurements obtained from qPCR and viral culture.

### Mathematical model

We used an empirical mathematical model describing viral kinetics for parameter inference [40]. The model is given by:

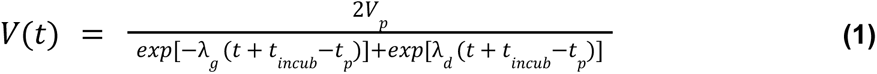

There are five parameters in this model: *V*_*p*_ represents the peak viral load, and *λ_g_* and *λ_d_* are the growth and decay rate of viral load, respectively;*t_incub_* represents incubation time for the pediatric and elderly group (data from observation studies), and we assume*t_incub_* = 0 for the adult group (data from a human challenge study); *t*_*p*_ is the time when viral load reaches a peak. Note that the model does not capture any underlying biological mechanisms underpinning viral infection, such as the host immune response, and therefore cannot be used to characterize every aspect of *in vivo* viral kinetics individually. Regardless, the model can provide insights into important and measurable characteristics of *in vivo* viral dynamic curves [41]. For example, the decay rate of viral load (*λ_d_* ) implicitly includes the effects of the immune response, indicating the average rate at which viral load decreases during the infection.

### Statistical inference

We applied a hierarchical Bayesian inference method to fit the mathematical model (**Eq. (1)**) to the virological data from the 53 individuals simultaneously. Here, five parameters were estimated (the parameter space: Φ = (*V_p_* , *λ_g_* , *λ_d_* ,*t_incub_* , *t*_*p*_ )) at both the age-group (i.e. study) and the individual levels. For each individual *i* in age group *a*, the parameter space Φ_*i,a*_ represented the individual-level parameters, and the predicted viral load values were given by *V*_*i,a*_ (*t*) = *f*(Φ_*i,a*_ ) + ɛ_*i,a*_ , where *f*_*i,a*_ was **Eq. (1)** and ɛ represented measurement errors for the individual *i* in age group *a*. The prior on the measurement error parameter was assumed to be normally distributed with mean 0 and variance 1. As we fitted the viral load data on a log10 scale, it was reasonable to set the variance to 1 log10 PFUe/ml. The individual-level parameters within the same age group were assumed to be drawn from age-specific distributions. For example, samples of peak viral load for each individual *i* in age group *a* were drawn from a normal distribution, 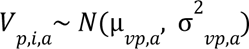 where µ_*vp,a*_ and 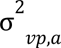 were the mean and variance of the peak viral load for age group *a*, respectively. Other parameters were modeled similarly. The age group-level parameters, for instance, µ_*vp,a*_ and 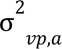 were given hyperpriors to reflect our prior beliefs about their values, and we assumed the same priors of each parameter for all age groups. Here, we assumed weakly informative priors (e.g., a distribution with large variance) for the mean of each group-level parameter. Based on previous estimates [12] and observational studies [28], we set µ_*vp,a*_ ∼ *N*(0, 3), µ_*tp,a*_ ∼ *N*(0, 3), µ_*t*_*incub,a*_ ∼ *N*(0, 3) *µ_λg,a_* ∼ *N*(1, 3), and *µ_λd,a_* ∼ *N*(1, 3). The variance of the distribution was 3, indicating that approximately 95% of the data falls within two standard deviations of the mean value (e.g., ±3.5 log10 PFUe/ml), which is reasonably large considering that the viral load of RSV kinetics ranged between 2 to 8 log PFUe/ml across different age groups. The variances of the group-level parameters were also weakly informative, such that all variances were assumed to follow a Cauchy distribution centered at 0 with a scale parameter of 2, indicating a variation of 2 log10 PFUe/ml.

Model fitting was performed in R (version 4.0.2) and Stan (Rstan 2.21.0). Samples were drawn from the joint posterior distribution of the model parameters using Hamiltonian Monte Carlo (HMC) optimized by the No-U-Turn Sampler (NUTS) (see [42] for details). In particular, we used five chains with different starting points and ran 5,000 iterations for each chain. The first 2,500 iterations were discarded as burn-in, and we retained 12,500 samples in total from the five chains.

### Estimation of infection probability

To estimate the probability of infectious virus establishing infection (*p_v_* ), we first defined *P_inf_* as the probability of one or more virions successfully establishing an infection in a recipient during viral shedding periods. The probability of a contact becoming infected during the viral shedding period (*P_inf_* ) is a function of the probability that a single viral particle infects the contact and the number of viral particles per time point during the infection 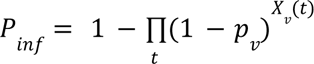. We computed the area under the viral load curve (*AUC_V_* ) during shedding periods (i.e., the cumulative viral load during viral shedding periods) based on the 12,500 population-level posterior samples for each age group. The value is given by

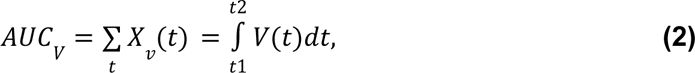

where *t*1 and *t*2 define the start and the end of shedding periods. We then estimated *p_v_* by assuming that *P_inf_* is equal to the empirical estimate of the secondary attack rate (SAR) from household transmission studies based on the SAR for pediatric index cases [18–20].

To validate the hypothesis that differences in the viral kinetics between age groups can explain differences in the risk of transmission, we then calculated *P_inf_* for the adult and elderly age groups based on the estimated value of *p_v_* for the pediatric age group and the values of *AUC_V_* estimated from the viral kinetic data. However, to account for differences in respiratory droplet spread between pediatric individuals and other groups, we estimated an additional parameter ξ based on the SAR for the adult group from one study [18]. Based on the equation 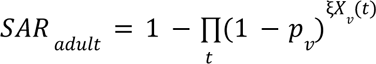, we calculate the parameter ξ, where *p_v_* is the median estimate based on SAR for pediatric individuals. We then used this value of ξ and the study-specific value of *p_v_* to predict the SAR for adult and elderly index cases in the other two studies [19,20].

### Model prediction

Based on the estimated posterior samples, we predicted the fraction of presymptomatic transmission based on the formula:

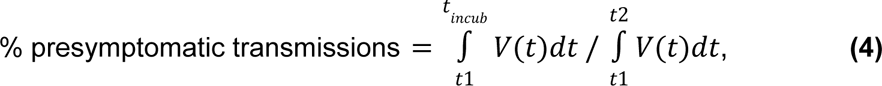

where *t*1 and *t*2 define the start and end of the shedding period, and*t_incub_* is the estimated incubation period. *V*(*t*) represents the predicted viral load time-series given by the posterior samples. The area under the viral load curve was calculated using *trapz* function in R (version 4.0.2). The method was also used in [8].

## Data Availability

All data produced in the present work are contained in the manuscript.

## Acknowledgments

This work was supported by a grant from the National Institutes of Health (R01AI137093). The content is solely the responsibility of the authors and does not necessarily represent the official views of the National Institutes of Health.

## Data and code availability

We used the R statistical software (v4.0.2) for all statistical analyses and visualization. Data and code used in this study are publicly available on Github: https://github.com/keli5734/RSV_Viral_Dynamics_Study

## Supplementary Figures

**Supplements Figure 1.**
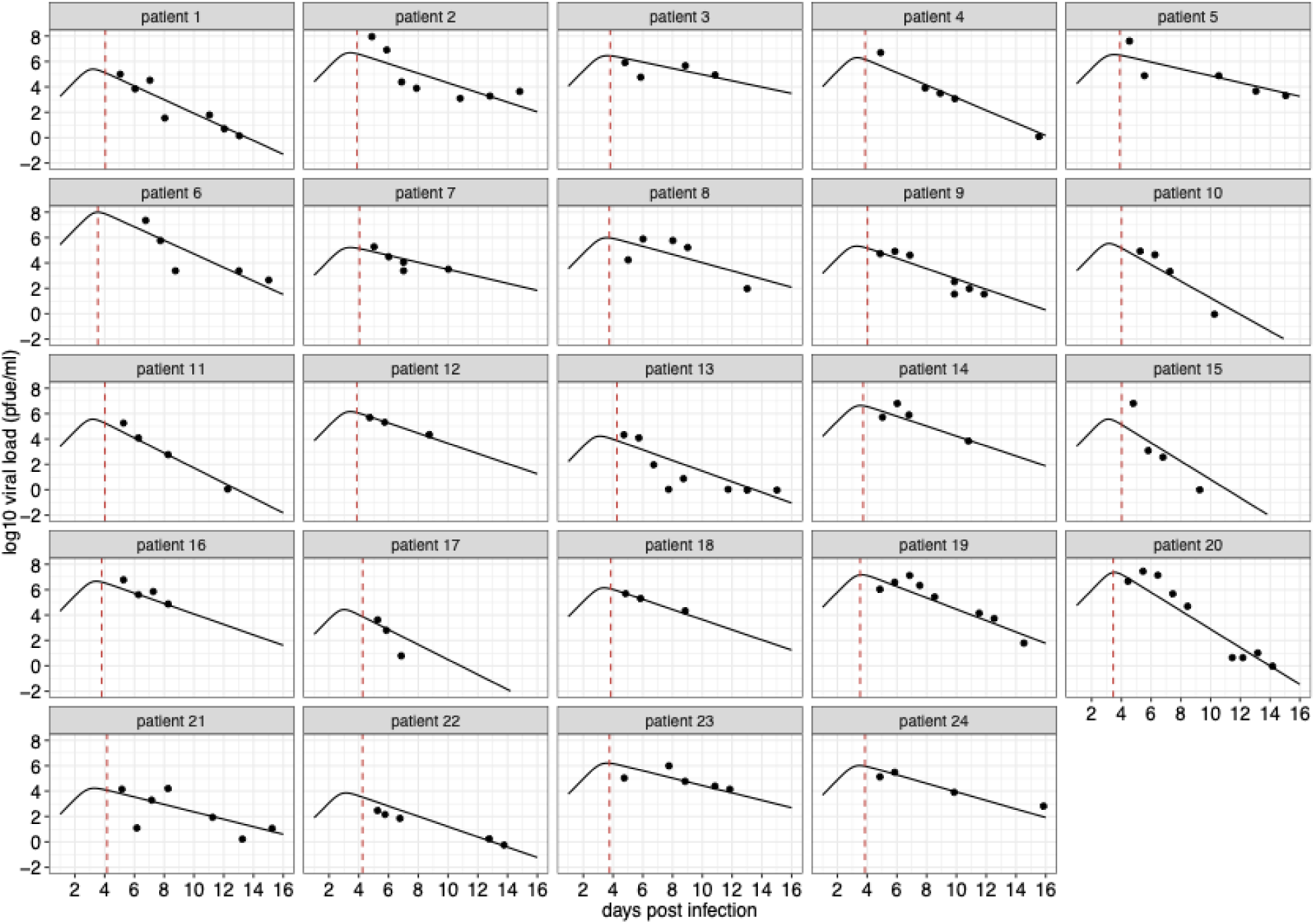
Model fits to viral kinetic data of RSV infections in the pediatric group. Data are presented with solid circles, and solid lines are median predictions of viral load. The dashed red lines indicate the median estimates of the time of symptom onset. Note that the median predictions of viral load are calculated using estimated parameters for each individual. Viral load data of patient 1-16 were from [14], and the data of patient 17-24 were from [13].

**Supplements Figure 2.**
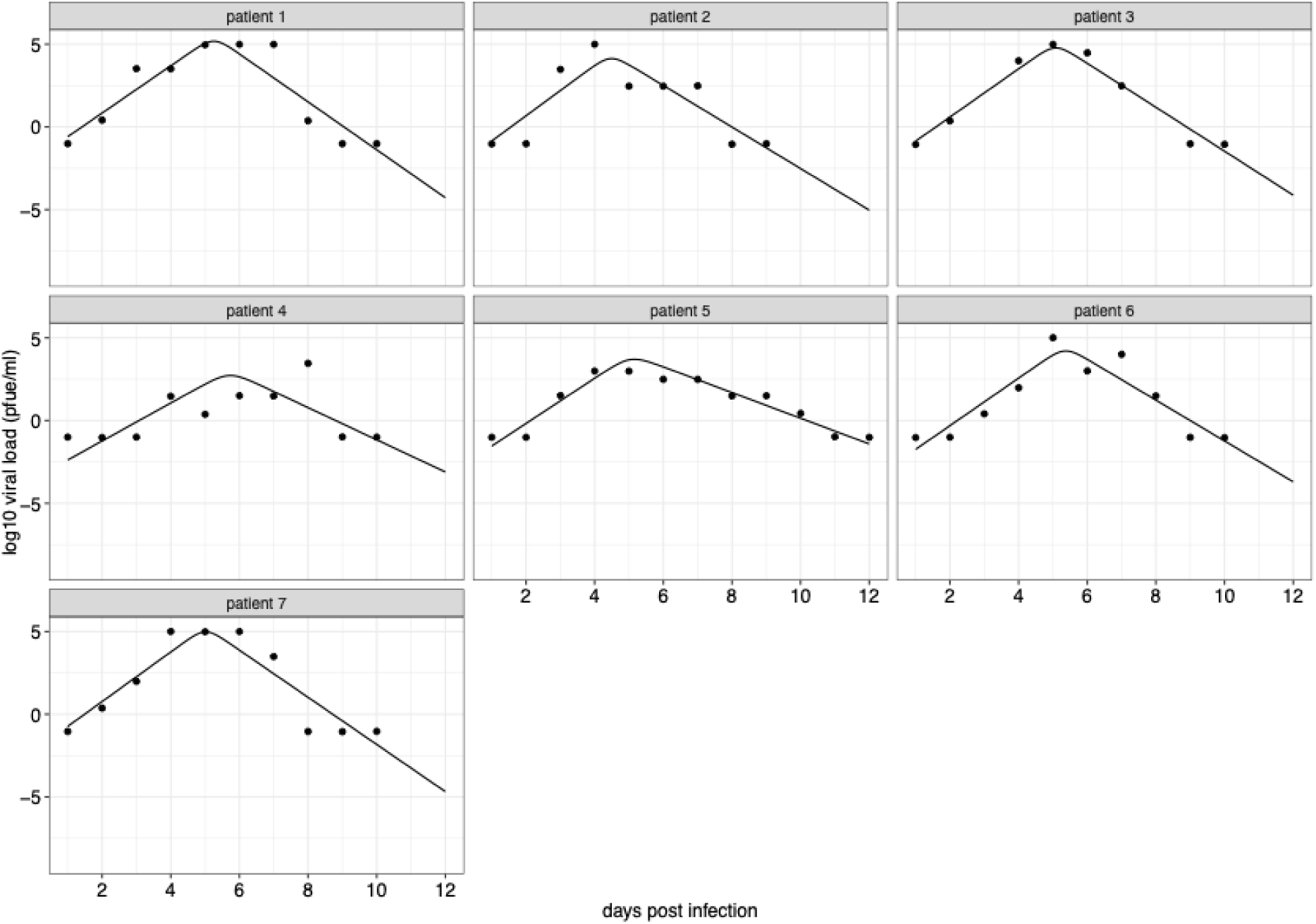
Model fits to viral kinetic data of RSV infections in the adult group. Data are presented with solid circles, and solid lines are median predictions of viral load. Note that the median predictions of viral load are calculated using estimated parameters for each individual.

**Supplements Figure 3.**
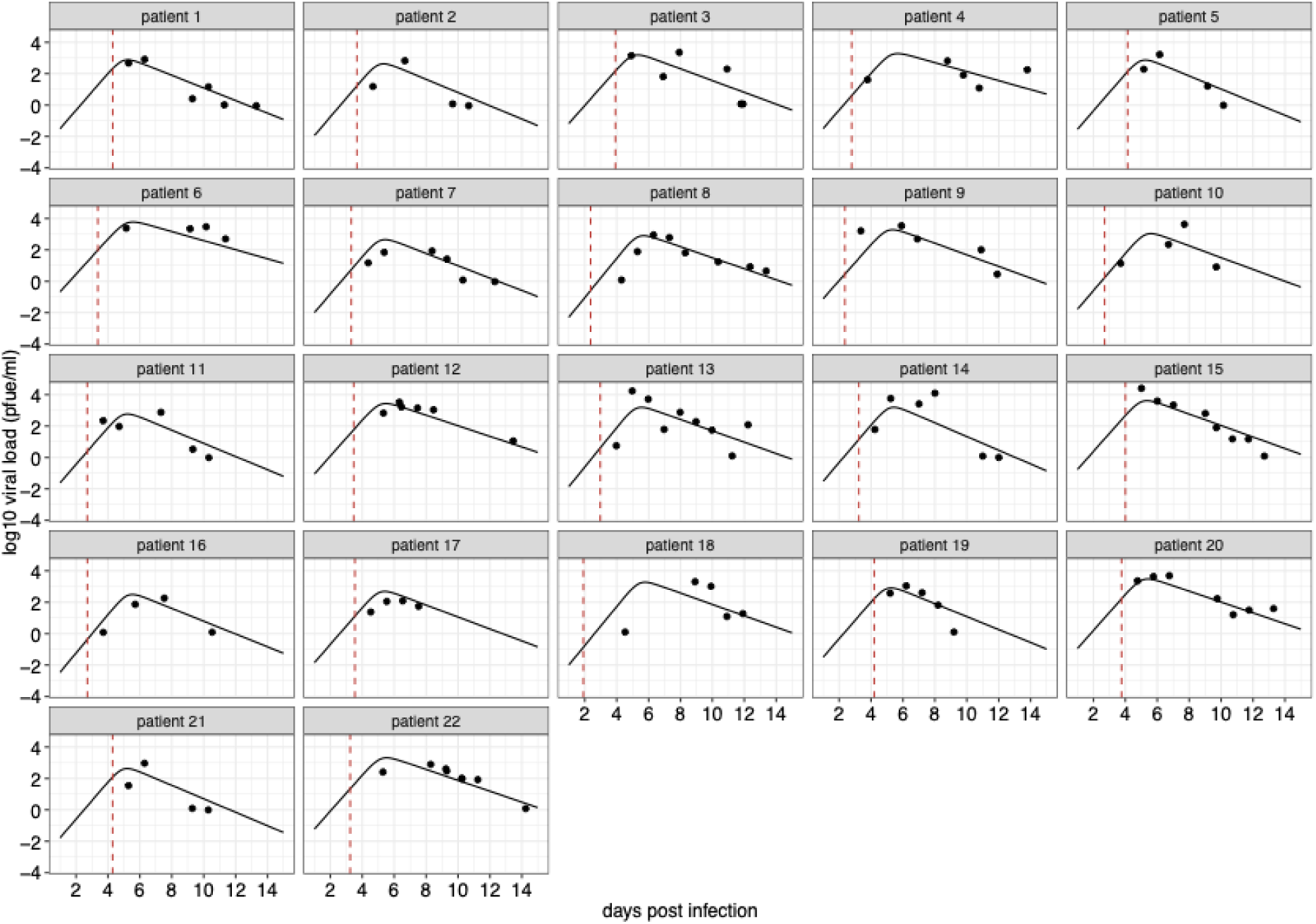
Model fits to viral kinetic data of RSV infections in the elderly group. Data are presented with solid circles, and solid lines are median predictions of viral load. The dashed red lines indicate the median estimates of the time of symptom onset. Note that the median predictions of viral load are calculated using estimated parameters for each individual.

**Supplements Figure 4.**
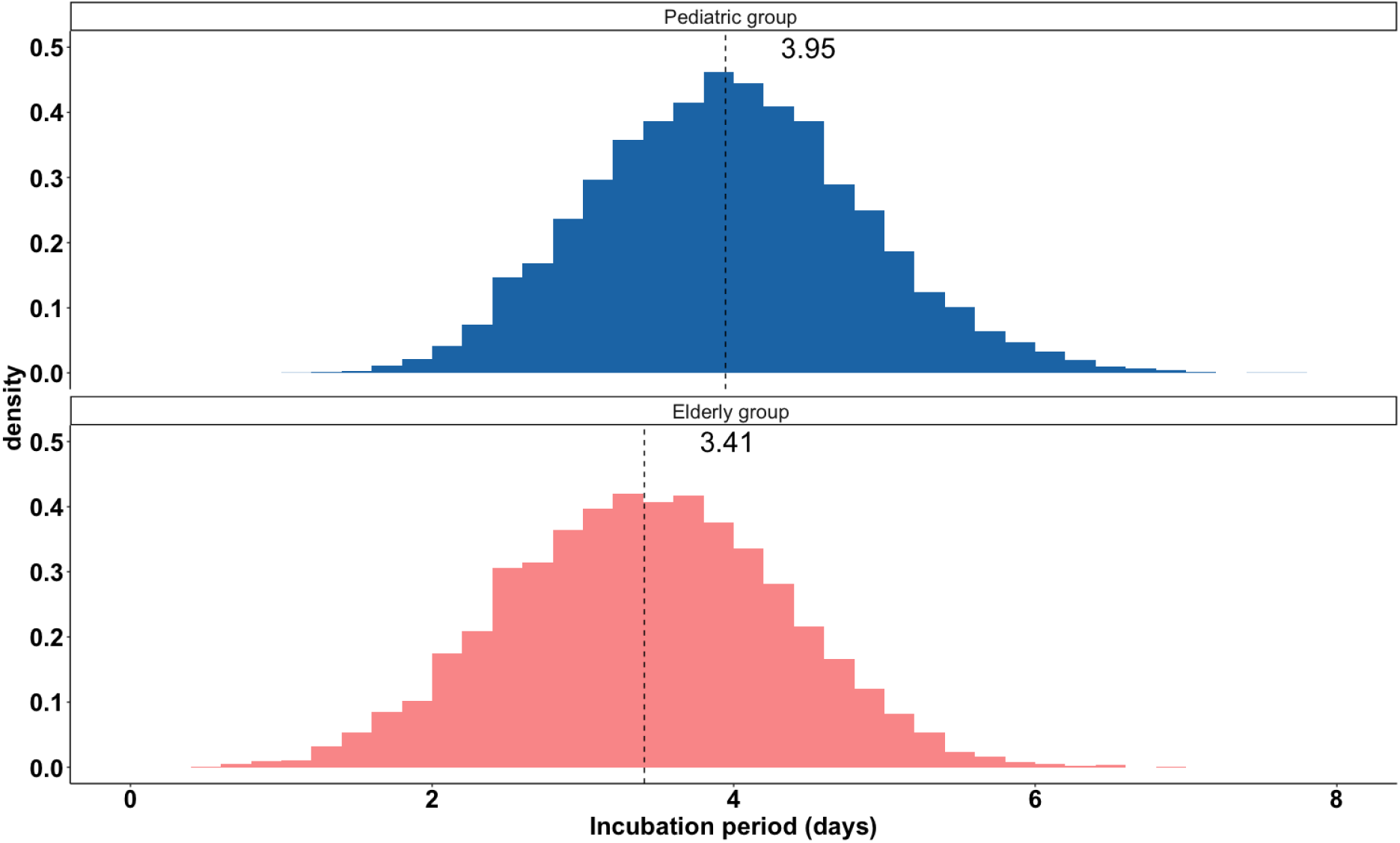
Posteriors distributions of incubation periods. 12,500 samples are drawn from the posterior distributions of incubation periods for the pediatric group (the upper panel) and the elderly group (the lower panel). Dashed lines indicate the median estimates.

**Supplements Figure 5.**
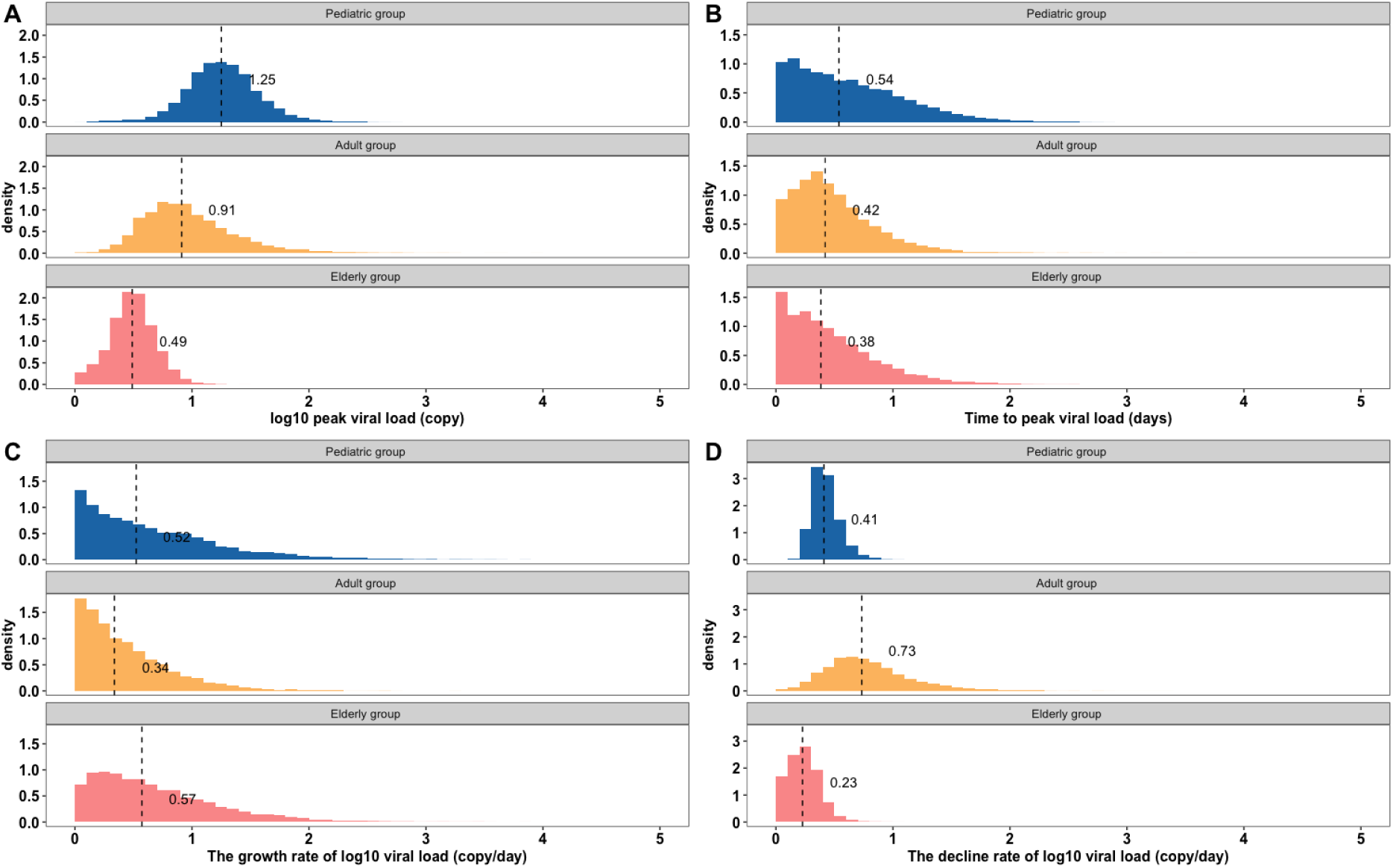
Posterior distributions showing the level of uncertainty (posterior standard deviations) of estimated model parameters. 12,500 samples are drawn from the posterior distributions of (A) peak viral load, (B) time to peak viral load, (C) the growth rate of viral load, and (D) the decline rate of viral load in the three age groups. Dashed lines indicated the median value of the posterior distributions.

**Supplements Figure 6.**
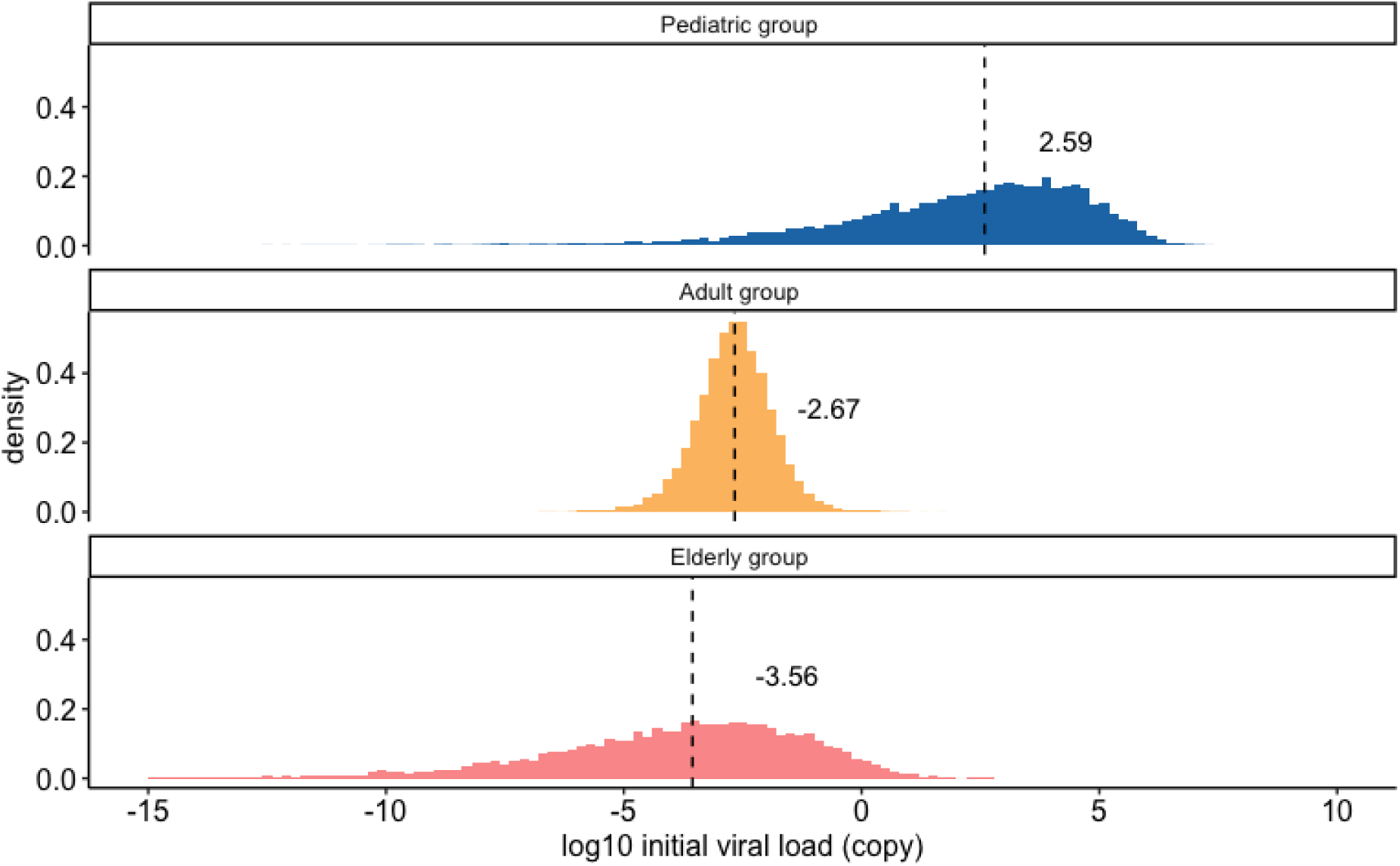
Distributions of initial viral load based on the population-level parameters. 12,500 samples are drawn from the posterior distributions to calculate the initial viral load for the pediatric group (the upper panel), the adult group (middle panel), and the elderly group (the lower panel). Dashed lines indicate the median estimates.

**Supplements Figure 7.**
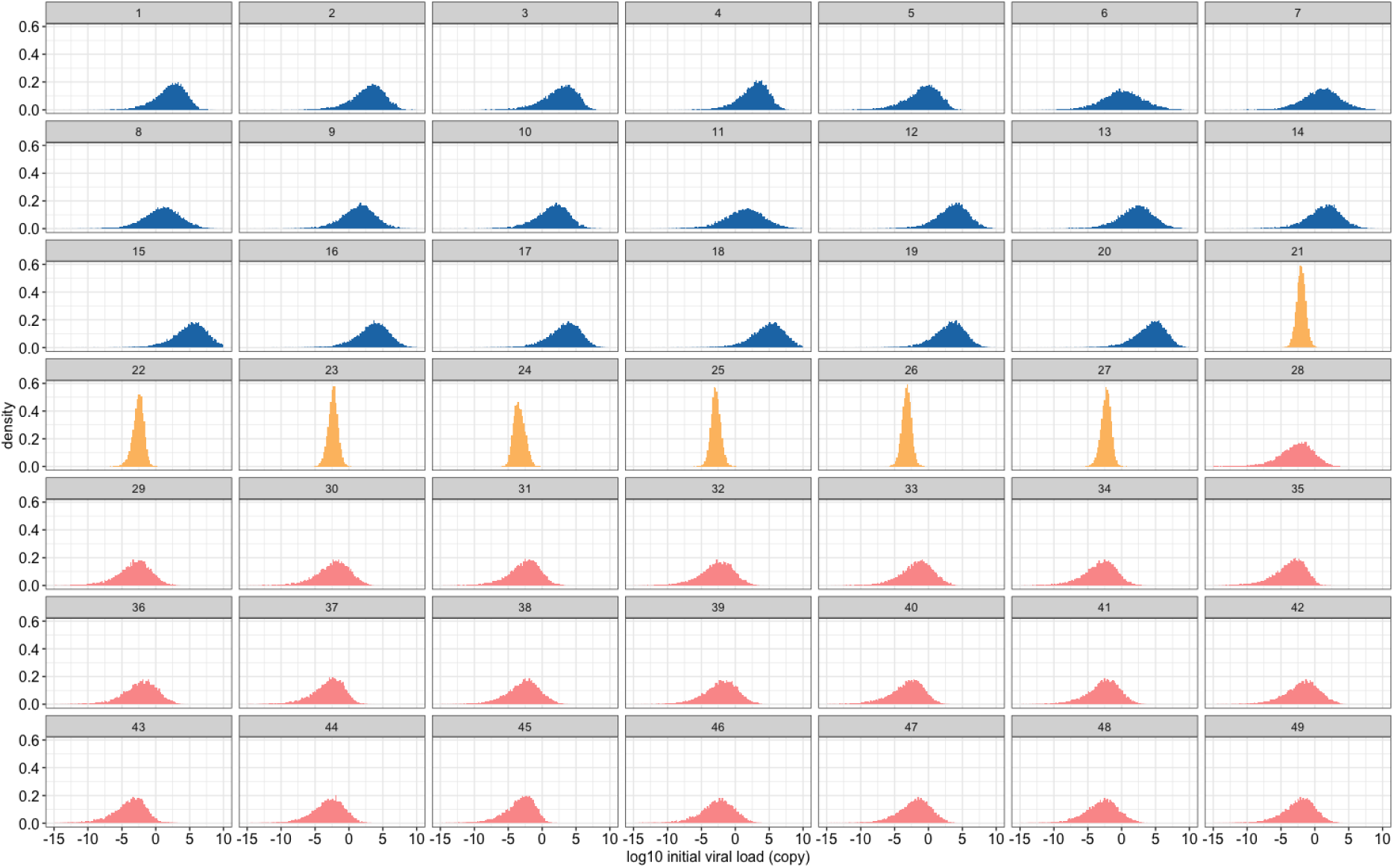
Distributions of initial viral load based on individual-level posteriors. 12,500 samples are drawn from the posterior distributions to calculate the initial viral load for the pediatric group (blue), the adult group (yellow), and the elderly group (red).

**Supplements Figure 8.**
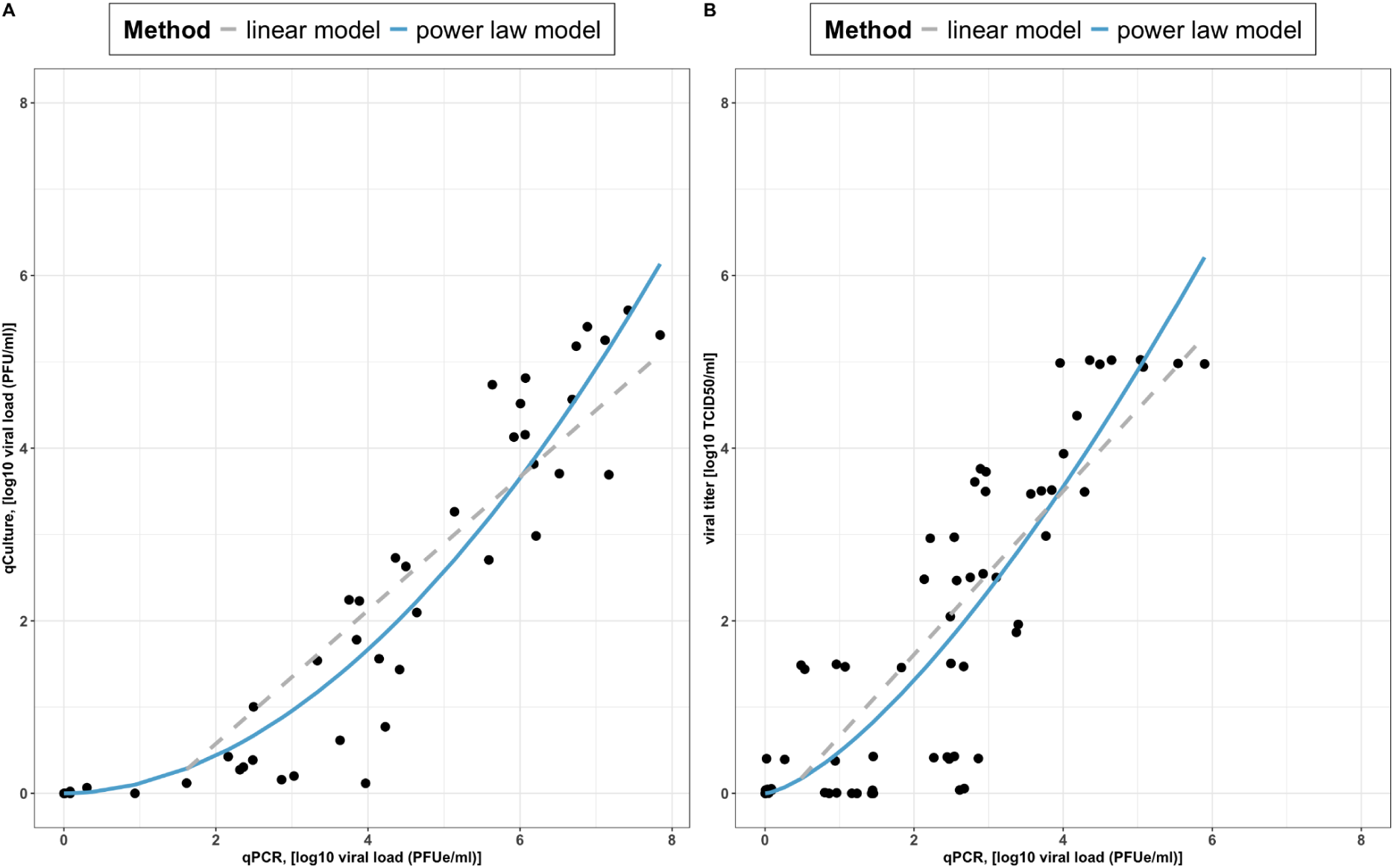
Characterization of total viral load with cell culture infectivity and transmission probability. **(A-B)** The model fits of using linear (in a form of *qCulture*(*t*) = *a* + *bV*(*t*), where *V*(*t*) is viral laid measured by qPCR) or power-law models (*qCulture*(*t*) = *cV^h^*(*t*)) to the viral loads measured by qPCR (horizontal axis) and cell culture (vertical axis) are based on the data from DeVincenzo et al. (*a* = –0. 97, *b* = 0. 77, *c* = 0. 11, *h* = 1. 93 ) and Falsey et al. (*a* = 0. 28, *b* = 0. 95, *c* = 0. 48, *h* = 1. 43 ), respectively.

**Supplements Figure 9.**
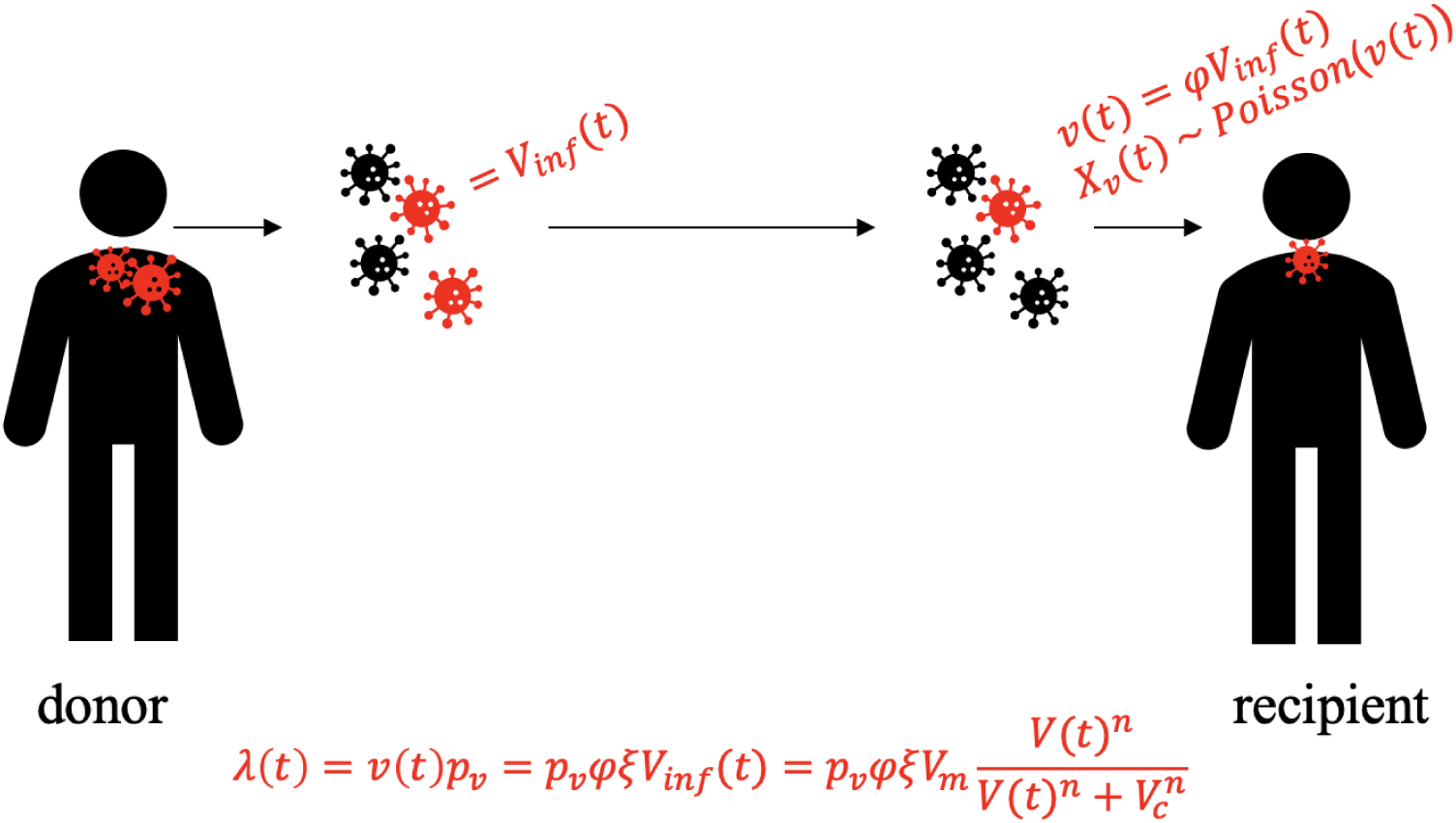
Illustrative model for transmission probability. To estimate the likelihood of transmission during a short contact duration τ, we assumed the total number of infectious viruses at time *t* is *V*_*inf*_ (*t*), and a proportion (φ) of of the viruses are transmitted to the recipient, such that *v*(*t*) = φ*V*_*inf*_(*t*). We also assumed that the number of infectious viruses reaching the recipient during a contact at time *t* is a random variable *X_v_*(*t*) that is Poisson distributed with the parameter *v*(*t*). We further assumed that each infectious virus has a probability *p_v_* to establish infection in a recipient. Since *X_v_* follows a Poisson distribution, we can show that the distribution of the number of viruses that successfully establish an infection follows a Poisson distribution with parameter λ = *v*(*t*)*p_v_* = φ*p_v_*ξ*V*_*inf*_(*t*) = *p_v_*ξφ*f*(*V*(*t*)), where *f* is a saturation function characterizing the relationship between total viral load and infectious viral load. The parameter ξ is a parameter that accounts for the number of infectious particles derived from the measured viral concentration (i.e., PFU/ml). We assumed the proportion φ = 1 in our analysis.

**Supplements Figure 10.**
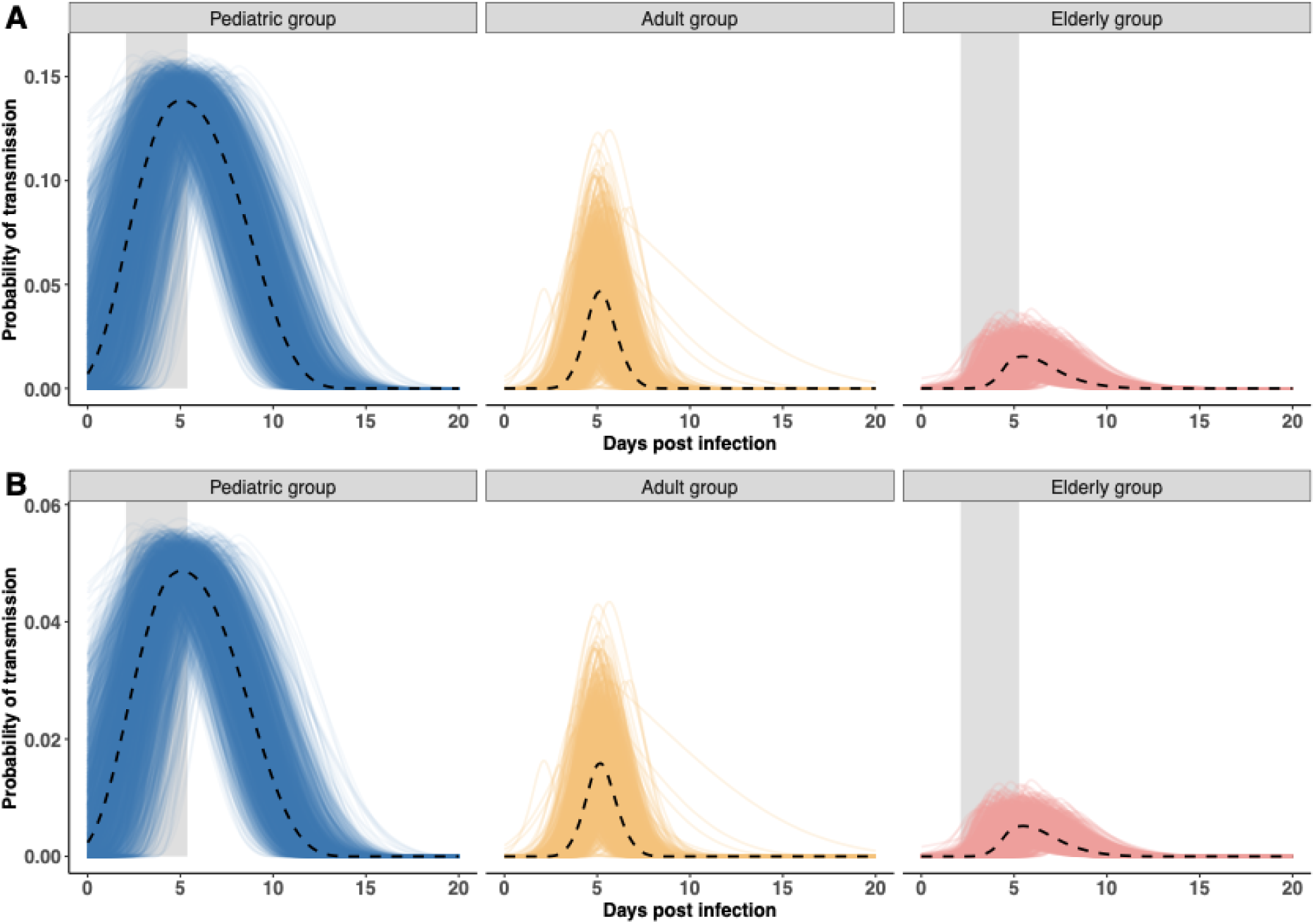
Probability of transmission in different age groups. 12,500 population-level samples are drawn from the posterior distribution to calculate the probability of transmission as time-series in different age groups, given the median estimates of infection probability from the household transmission studies **(A)** [20] and **(B)** [18]. Dashed lines indicate the median trajectory, and shaded areas indicate estimated incubation period in the pediatric and elderly group.

